# Association between Mindful Eating and Obesity, Nutritional Status among 6th Grade Medical School Students

**DOI:** 10.1101/2025.04.29.25326508

**Authors:** Penbe Ecem Misirlioğlu, Hatice Şimşek

## Abstract

**Purpose:** This study aimed to examine the association between mindful eating, obesity, and nutritional status (low diet quality) among 6th year medical students of the Dokuz Eylül University Medical School.

**Material and Methods:** The population of this cross-sectional study consisted of 303 interns studying at the Dokuz Eylül Medical School, for the year 2020-2021. No sampling method was applied; instead, the entire population was targeted. The dependent variable was presence of obesity determined by the body mass index and low diet quality determined by the Mediterranean Diet Quality Scale (KIDMED). The Independent variable was mindful eating determined by the Mindful Eating Questionnaire (MEQ-30). Possible confounding factors include socio-demografic characteristics, health status, dietary behaviour, the presence of obesity in the family, physical activity status. The data were collected through an online survey form. The percentage distributions of the descriptive variables were presented with mean±standard deviations, Chi-Square and Multiple Logistic Regression Analysis were used in order to determine casuality.

**Results:** The mean age of 245 interns reached was 23.9±1.2 (range 22-30), 42.9% of whom were women. The mean total YFO-30 scores were 98.3±11.3. The frequency of obesity was 4.1%. In this study, 44.9% of the students had low diet quality. According to the Logistic Regression Analysis the risk of being overweight or obese was significantly higher 3.4 (OR:3.44 [95%CI:1.06-11.16] p= 0.039) times in those aged 26 and over, 10.7 (OR:10.70 [95%CI:4.50-25.47] p<0.001) times in males, 2.5 (OR:2.53 [95%CI: 1.27-5.03] p=0.008) times in those with obese individuals in the family; the risk of low diet quality is meaningfully higher 2,1 (OR:2.08 [95%CI:1.16-3.74] p=0.014) times in males, 3.0 (OR:2.95 [95%CI:1.22-7.17] p=0.005) times those whose income is lower than their expenses, 3.1 (OR:3.12 [95%CI:1.42-6.87] p=0.005) times in those that are living with their friends.

**Conclusion:** Frequency of obesity was low, low diet quality was high in 6th grade medical students. There was no significant relationship between mindful eating and low diet quality and obesity.

## INTRODUCTION

Unhealthy eating habits and obesity among university students pose critical public health challenges. Studies have found the prevalence of obesity in university students to be 2.1-15.5% (1). The causes of obesity in university students are generally stated as skipping meals, especially breakfast, frequent consumption of foods such as bagels and tea, lack of time for healthy nutrition and lack of healthy nutrition knowledge (1-2).

University students often begin living independently for the first time, which significantly influences their eating habits. The literature indicates that many young adults lack the necessary nutrition education and experience to make healthy dietary choices. Irregular and demanding academic schedules, coupled with limited food preparation skills, further complicate the development of balanced eating patterns. Additionally, students frequently encounter emotional challenges, inadequate nutritional knowledge, and socioeconomic constraints, all of which contribute to unhealthy eating behaviors (3).

University life may therefore facilitate the adoption of unfavorable dietary and lifestyle practices, which are likely to persist into adulthood and adversely affect long-term health outcomes. Accordingly, for the health of future generations, it is crucial to investigate the complex and interrelated dynamics among food, nutrition, and individual behavior through a multidimensional and evidence-based approach. In recent years, mindful eating has gained attention as a behavioral intervention aimed at promoting healthier dietary practices. Mindful eating refers to the conscious awareness of internal experiences both emotional (e.g. stress, anger, anxiety) and physiological (e.g., hunger and satiety cues) during food consumption (4). This approach enables individuals to attenuate the impact of emotional states on eating behavior, thereby reducing maladaptive patterns such as emotional eating and automatic or habitual eating. Moreover, mindful eating has been associated with improved regulation of energy intake and enhanced dietary self-control (5). Evidence suggests that this practice may contribute to weight reduction in individuals with obesity by decreasing the pace of eating and mitigating non-homeostatic, stress-induced food intake. Binge eating, emotional eating, external eating, and eating in response to food cravings are also effective in weight regain after successful weight loss (6). Studies have found a negative relationship between mindful eating and Body Mass Index (BMI) in young adults (6-8). Mindful eating helps to gradually transform extrinsic motives for eating into intrinsic motives and promotes healthier eating behaviors such as increased intake of fruits and vegetables and decreased consumption of high-sugar and energy-dense foods (9-10).

It is crucial for healthcare professionals, who have an important role in protecting and improving the health of society, to be aware of their own eating habits and eating attitudes from their student years in terms of maintaining body weight and adopting healthy eating habits (11). For these reasons, this study was conducted to determine the relationship between mindful eating and obesity and nutritional status (low diet quality) in 6th-semester students at Dokuz Eylül Faculty of Medicine.

## MATERIALS AND METHODS

The population of this cross-sectional study consisted of all 6th-semester students (n = 303) enrolled at Dokuz Eylül University Faculty of Medicine during the 2020–2021 academic year. No sampling was performed, as the study aimed to include the entire population. The dependent variables were the presence of obesity and poor diet quality. Obesity was determined based on body mass index (BMI), which was calculated using self-reported height and weight. Those with a BMI of <18.5 kg/m^2^ were classified as underweight; those with a BMI of 18.50-24.99 kg/m^2^ as normal weight; those with a BMI of 25.0-29.99 kg/m^2^ as overweight; and those with a BMI of >30.0 kg/m^2^ as obese (12). Nutritional status was determined according to the Mediterranean Diet Quality Index (KIDMED), which was developed by Serra Majem et al. in 2004 (13). The Turkish validity and reliability were assessed by Akar Şahingöz et al. The scale consists of 16 items with “yes” and “no” options. Of the questions included in KIDMED, 12 are positive questions, and 4 are negative questions. If the total score is ≥8, it is interpreted as “good Mediterranean diet”; if it is between 4-7, it is interpreted as “the suitability of the Mediterranean diet should be improved (medium)”; and if it is ≤3, it is interpreted as “very low diet quality (low)” (14). The independent variable is the level of mindful eating as determined by the Mindful

Eating Questionnaire (MEQ-30). The Mindful Eating Questionnaire (MEQ), which consists of 28 items developed by Framson et al. was used to determine the level of eating mindfulness. Individuals with high scores on this scale are considered to be aware of and respond to physiological indicators of hunger and satiety (3). The Turkish validity and reliability of the scale were studied by Köse et al. (2016). The total mindful eating score is determined by the sum of the scores between 1-5 given to each question (taking into consideration the scores of the reverse questions) (15). Possible confounding factors include sociodemographic characteristics, health status, nutritional behaviors, the presence of obesity in the family, and physical activity status. In the study, 245 students were reached (response rate: 80.9%). Data were collected using a “Google survey” form, which was sent via two phone messages through intern representatives from each internship group. The SPSS 24.0 software package was used for data analysis. Descriptive data were presented as the number and percentage distributions for categorical variables and as mean ± standard deviation for continuous variables. In causal analyses, Chi-square analysis, t-tests, and multiple logistic regression analysis (as multivariate analysis) were used to determine the relationship between dependent and independent variables. In univariate analyses, the variables that showed a significant relationship with the dependent variable were further analyzed using logistic regression analysis.

### Ethical Considerations

Ethics Committee approval for the study was obtained from the Dokuz Eylül University Faculty of Medicine, Non-Interventional Studies Ethics Committee (2020/26-35). The study was conducted in accordance with the Declaration of Helsinki, and all participants provided informed consent at the beginning of the web-based questionnaire.

## RESULTS

The mean age of the 245 intern physicians was 23.9 ± 1.2 years (range: 22-30), and 42.9% (n = 105) were female. Of the students, 79.8% lived with friends, 37.2% of the students’ mothers and 51.4% of the students’ fathers had a university education, and 52.3% of the students’ family income matched their expenses. 10.6% of the participants had at least one chronic disease (n = 26), with endocrine system diseases (46.1%), cardiovascular diseases (14.5%), digestive system diseases (11.5%), and hypertension (7.6%) being the most common, in that order. According to Table 1, 18.8% of the students smoked, 66.9% skipped meals, and the most frequently skipped meal was breakfast (58.6%). Most of them consumed junk food between meals (71.4%). 32.3% of the participants consumed fast food two or more times a week. 4.1% of the interns were obese, 25.7% were overweight, and 44.9% had low diet quality. The mean total score of the MEQ-30 was 98.3 ± 11.3 (range: 68-133). Descriptive findings are shown in Table 1.

**Table 1.**
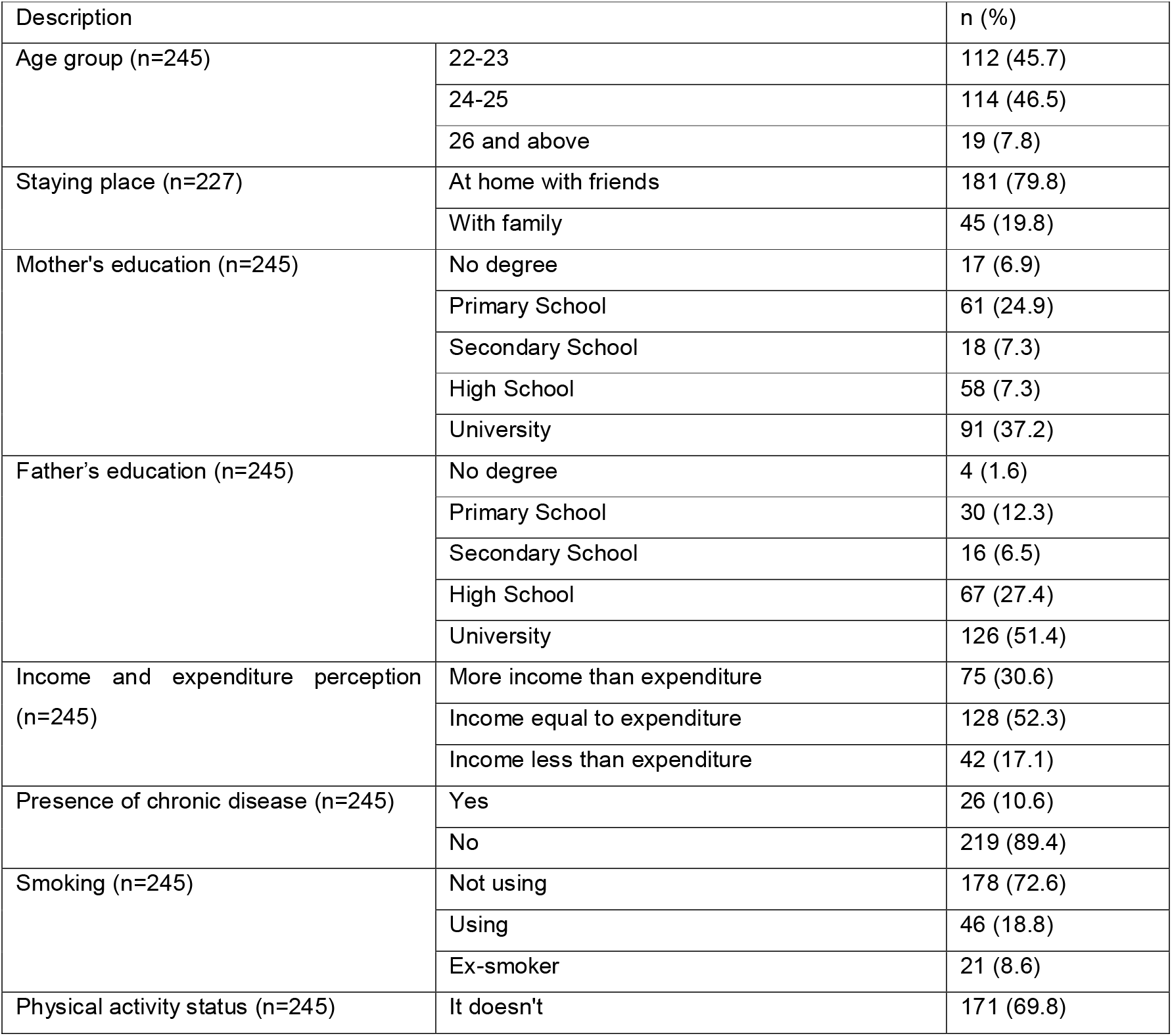
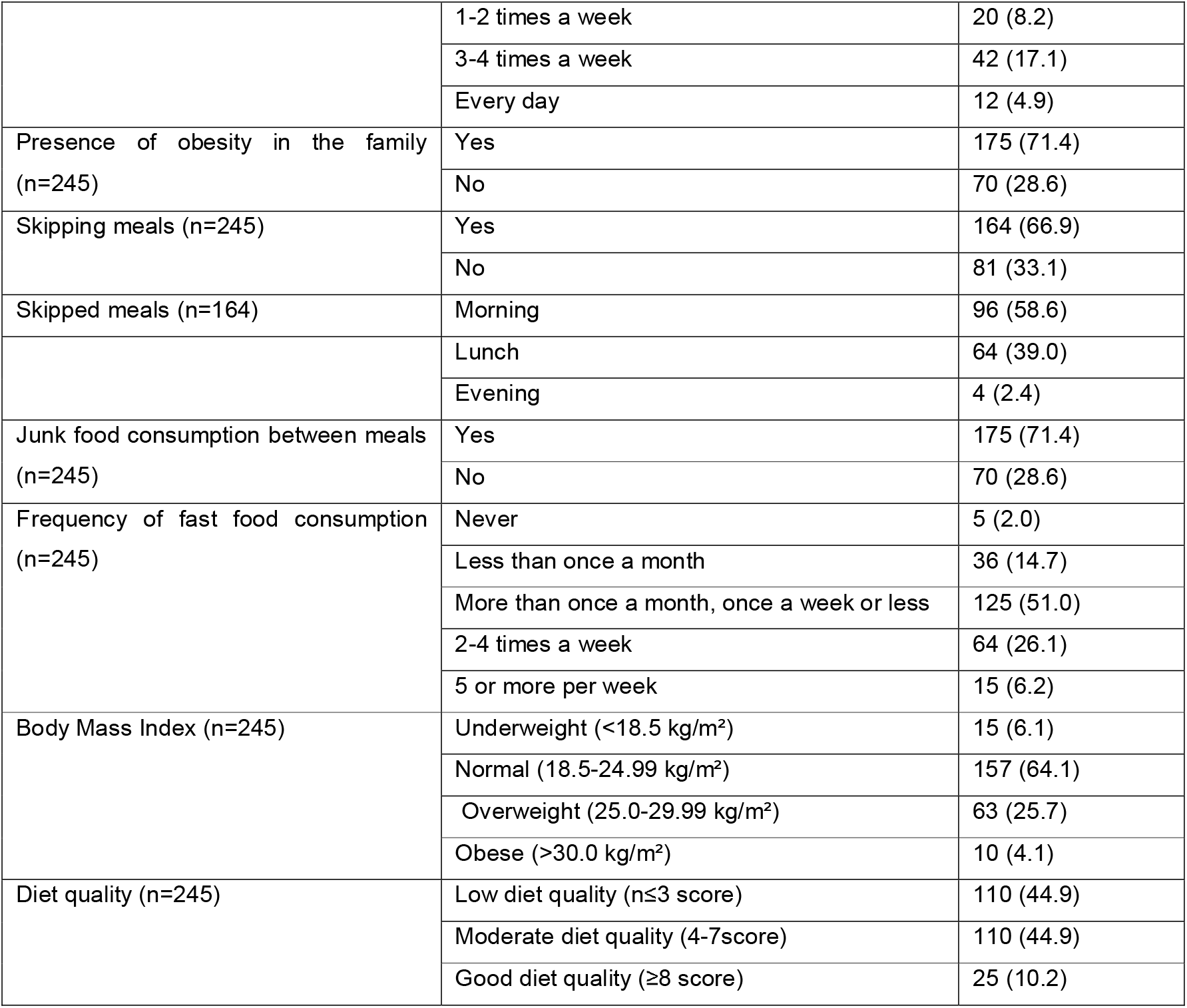
Distribution of participants according to their descriptive characteristics.

In univariate analyses, while being overweight-obese increased significantly with increasing age group (p=0.018), being overweight-obese was significantly higher in males than females (p<0.001). In addition, the presence of obesity in the family (p=0.012) and smoking (p=0.002) were significantly associated with being overweight-obese (Table 2). According to univariate analyses, diet quality was significantly lower in males (p=0.004), those whose mothers had primary school education or less (p=0.044), and those who stayed at home with friends (p<0.001). According to income and expenditure perception data, the lower the family income, the significantly higher the poorer the diet quality (p=0.003) (Table 2).

**Table 2.** Association of students’ socio-demographic characteristics with poor diet quality and being overweight-obese.

| Description | Being overweight-obese |  | Low diet quality |  |
| --- | --- | --- | --- | --- |
|  | n (%) | p | n (%) | P |
| Age group |  | **0.018\*** |  | 0.851 |
| 22-23 (n=112) | 24 (21.4) |  | 52 (46.4) |  |
| 24-25 (n=114) | 40 (35.1) |  | 49 (43.0) |  |
| 26 and above (n=19) | 9 (47.4) |  | 9 (47.4) |  |
| Gender |  | **<0.001** |  | **0.004** |
| Woman (n= 105) | 9 (8.6) |  | 36 (34.3) |  |
| Men (n= 140) | 64 (45.7) |  | 74 (52.9) |  |
| Mother's education (n=245) |  | 0.308* |  | **0.044\*** |
| Primary school and below (n= 78) | 27 (34.6) |  | 44 (56.4) |  |
| Secondary and high school (n=76) | 24 (31.6) |  | 29 (38.2) |  |
| University and higher (n=91) | 22 (24.2) |  | 37 (40.7) |  |
| Father's education |  | 0.074* |  | 0.106* |
| Primary school and below (n=36) | 15 (41.7) |  | 22 (61.1) |  |
| Secondary and high school (n=83) | 28 (33.7) |  | 35 (42.2) |  |
| University and higher (n=126) | 30 (23.8) |  | 53 (42.1) |  |
| Perception of income and expenditure |  | 0.245* |  | **0.003\*** |
| More income than expenditure (n=75) | 20 (26.7) |  | 24 (32.0) |  |
| Income equal to expenditure (n=128) | 36 (28.1) |  | 59 (46.1) |  |
| Income less than expenditure (n=42) | 17 (40.5) |  | 27 (64.3) |  |
| Place of stay |  | 0.878 |  | **<0.001** |
| With family (n=46) | 14 (30.4) |  | 11 (23.9) |  |
| At home with friends (n=181) | 53 (29.3) |  | 95 (52.5) |  |
| Smoking |  | **0.002** |  | 0.096 |
| No (n=178) | 42 (23.6) |  | 73 (41.0) |  |
| Yes (n=46) | 22 (47.8) |  | 27 (58.7) |  |
| Ex-smoker (n=21) | 9 (42.9) |  | 10 (47.6) |  |
| Presence of chronic disease |  | 0.307 |  | 0.892 |
| Yes (n=26) | 10 (38.5) |  | 12 (46.2) |  |
| No (n=216) | 63 (28.8) |  | 98 (44.7) |  |
| Skipped meal |  | 0.649 |  |  |
| No skipping meals (n=81) | 24 (29.6) |  |  |  |
| Morning (n=96) | 26 (27.1) |  |  |  |
| Lunch, dinner (n=68) | 23 (33.8) |  |  |  |
| Junk food consumption |  | 0.508 |  |  |
| Yes (n=175) | 50 (28.6) |  |  |  |
| No (n=70) | 23 (32.9) |  |  |  |
| Frequency of fast food consumption |  | 0.182 |  |  |
| Never/ once a week and less (n=166) | 45 (27.1) |  |  |  |
| 2 or more per week (n=79) | 28 (35.4) |  |  |  |
| Being physically active |  | 0.952 |  |  |
| Not doing regular physical activity (n=171) | 50 (29.2) |  |  |  |
| 1-2 days a week (n=20) | 6 (30.0) |  |  |  |
| 3 or more per week (n=54) | 17 (31.5) |  |  |  |
| Presence of obesity in the family |  | **0.012** |  |  |
| Yes (n=70) | 29 (41.4) |  |  |  |
| No (n=175) | 44 (25.1) |  |  |  |
| Level of diet quality |  | 0.725 |  |  |
| Low diet quality (n ≤ 3 score) (n=110) | 32 (29.1) |  |  |  |
| Moderate diet quality (4-7 score) (n=110) | 35 (31.8) |  |  |  |
| Good diet quality (≥ 8 score) (n=25) | 6 (24.0) |  |  |  |

\* Chi-Square Analysis in Slope

In the study, there was no significant difference in the mean scores of mindful eating between the overweight-obese group and the non-obese group and between the group with poor quality of life and the non-obese group (p=0.443; p=0.861) (Table 3). In the Logistic Regression model created with variables that were found to have a significant relationship with the dependent variable in univariate analyses in addition to the mindful eating score, no significant relationship was found between the mindful eating score and the risk of being overweight-obese (OR: 1.01, 95% CI: 0.99-1.04, p=0.209) and the risk of poor diet quality (OR: 0.95, 95% CI: 0.47-1.94, p=0.908) (Table 4, Table 5). However, the risk of being overweight-obese was 3,4 times (95% CI:1.06-11.16, p=0.039) higher in those aged 26 years and older, 10.7 times (95% CI:4.50-25.47, p<0.001) higher in men, and 2.5 times (95% CI:1.27-5.03, p=0.008) higher in those with obese family members. The risk of poor diet quality was 2.1 times (95% CI:1.16-3.74, p=0.014) higher in males, 3.0 times (95% CI:1.22-7.17, p=0.005) higher in those whose income was lower than expenses, and 3.1 times (95% CI:1.42-6.87, p=0.005) higher in those who stayed at home with friends.

**Table 3.**
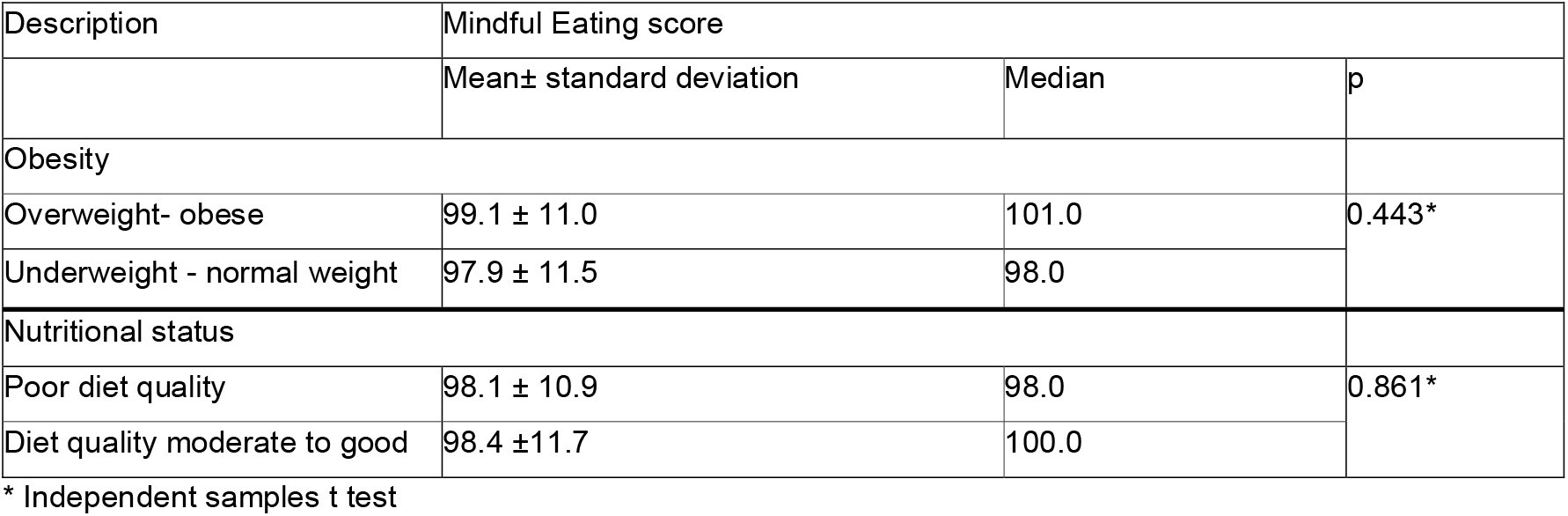
The relationship between students’ mean scores of eating awareness and diet quality and being overweight-obese.

| Description | Mindful Eating score |  |  |
| --- | --- | --- | --- |
|  | Mean± standard deviation | Median | p |
| Obesity |  |  | 0.443* |
| Overweight- obese | 99.1 ± 11.0 | 101.0 |  |
| Underweight - normal weight | 97.9 ± 11.5 | 98.0 |  |
| Nutritional status |  |  | 0.861* |
| Poor diet quality | 98.1 ± 10.9 | 98.0 |  |
| Diet quality moderate to good | 98.4 ±11.7 | 100.0 |  |

\* Independent samples t test

**Table 4.** Factors affecting the risk of being overweight-obese - Logistic Regression analysis results.

| Description (Reference group) |  | Being overweight-obese |  |  |
| --- | --- | --- | --- | --- |
|  |  | p | OR | 95% GA |
| Eating mindfulness total score (Continuous) |  | 0.209 | 1.01 | 0.99-1.04 |
| Age (22-23) | 24-25 | 0.120 | 1.69 | 0.87-3.28 |
|  | 26 years and older | **0.039** | **3.44** | **1.06-11.16** |
| Sex (Woman) | Man | **<0.001** | **10.70** | **4.50-25.47** |
| Smoking (Does not use) | Ex smoker | 0.358 | 1.61 | 0.58-4.50 |
|  | Yes | 0.163 | 1.73 | 0.80-3.74 |
| Presence of obese individuals in the family (No) | Yes | **0.008** | **2.53** | **1.27-5.03** |

**Table 5.** Factors affecting poor diet quality - Logistic Regression analysis results.

| Description (Reference groups) |  | Low diet quality |  |  |
| --- | --- | --- | --- | --- |
|  |  | p | OR | 95% GA |
| Eating mindfulness total score (Continuous) |  | 0.908 | 0.95 | 0.47-1.94 |
| Sex (Woman) | Man | **0.014** | **2.08** | **1.16-3.74** |
| Mother's learning<br>(University and above) | Middle-high school | 0.343 | 0.70 | 0.34- 1.45 |
|  | Primary school and below | 0.291 | 1.46 | 0.72-2.94 |
| Place of stay (with family) | At home with friends | **0.005** | **3.12** | **1.42-6.87** |
| Perception of income and expenditure<br>(Income higher than expenditure) | Income equal to expenditure | 0.134 | 1.69 | 0.85-3.37 |
|  | Income lower than expenditure | **0.017** | **2.95** | **1.22-7.17** |

## DISCUSSION

In a study conducted among 6th-year students at Dokuz Eylül University Faculty of Medicine (2020– 2021), 44.5% of participants had low diet quality, 25.7% were overweight, and 4.1% were classified as obese. No statistically significant association was found between mindful eating and low diet quality or obesity. The average MEQ-30 score was 98.3, with female students scoring slightly higher than their male counterparts. Similar trends were reported in other Turkish studies (15–16).

The prevalence of obesity a recognized global public health concern is rising in both developed and developing nations. A multicenter study conducted among university students from various countries reported the prevalence of overweight and obesity as follows: Bangladesh (20.8%), China (2.9– 14.3%), Malaysia (20–30%), Thailand (31%), Pakistan (13–57.6%), India (11–37.5%), Colombia (12.4–16.7%), Mexico (31.6%), Kuwait (42%), Iran (12.4%), and Turkey (10–47%) (17).

University students often exhibit poor dietary habits, and in the present study, 79.8% of participants reported living with friends, a factor that may contribute to increased consumption of unhealthy foods. Although no significant relationship was found between mindful eating and the risk of overweight or obesity, factors such as age, gender, and family history of obesity were found to influence this risk.

Previous studies on the relationship between mindful eating and BMI have shown mixed results, with some indicating a negative correlation (18).

The mean Mediterranean Diet Quality Index score in the current study was 3.9, with 44.9% of participants categorized as having low diet quality and only 10.2% as having good diet quality. Similar studies among university students in Turkey have reported good diet quality prevalence ranging from 2.3% to 17.8%, and poor diet quality ranging from 29.2% to 56.3% (19–21). These results suggest persistent suboptimal adherence to the Mediterranean diet among this population. Factors such as living away from family, lower socioeconomic status, and the high stress associated with medical education likely contribute to unhealthy eating behaviors.

A cross-sectional study using the KIDMED index among 354 medical students (206 first-year and 148 third-year) found that 19% of first-year and 31.3% of third-year students had poor adherence to the Mediterranean diet, suggesting a decline in diet quality with academic progression (22). For instance, a study in the United States reported that 34% of university students showed low, and 20% showed high, adherence to the Mediterranean diet (23). In Italy, only 20% of medical students were found to have poor diet quality, with male students displaying significantly lower adherence compared to female students (24). Additionally, a multicountry study applying the KIDMED index to 24-hour dietary recalls highlighted dietary patterns across five Mediterranean countries. Spain demonstrated the highest adherence to the Mediterranean diet, especially among adolescents, likely due to the cultural embedding of traditional Mediterranean food practices, strong family meal structures, and effective public health policies. In contrast, countries such as Croatia and Turkey despite their geographical proximity to the Mediterranean reported lower adherence levels, particularly among adolescents and young adults. These differences may stem from dietary Westernization, increased fast food intake, and lifestyle changes, indicating a need for targeted dietary interventions (25).

The Mediterranean diet is fundamentally characterized by a high intake of plant-based foods such as whole grains, fruits, vegetables, legumes, and nuts; moderate consumption of white meat, fish, eggs, and dairy products; low consumption of red and processed meats; and the use of olive oil as the primary fat source (19). The reason why students in Turkey mostly have low diet quality may be that many of them do not have a good nutritional pattern because they live separately from their families or face economic problems in accessing healthy food, which is also the result of our study. In addition, an important feature of studying in the final year of medical school in Turkey is the stress of the Medical Specialization Examination. As the level of education of medical students increases, they may not be able to spare time for healthy nutrition due to the intensity of courses and clinical practices. Due to this stress and intense work schedule, the diet quality of intern physicians may be low. No significant relationship was found between mindful eating scores and diet quality using the KIDMED index. Logistic regression indicated that being male, having lower income, and living with friends were linked to poorer diet quality. Living at home with friends may have increased consumption of nutrient-poor convenience foods such as fast food. Similar to our study, being male and having a low income level were found to negatively affect diet quality in university students. Previous studies have shown that diet quality may be low in university students with high income levels. Barbosa et al. (2016) among university students in São Luís, Brazil, showed that the prevalence of metabolic syndrome risk factors was higher among middle and high-income students. (26). This finding can be attributed to the impact of high income level on students lifestyle, for example, consuming fast food, eating an unhealthy diet high in fat and high in carbohydrates, eating between meals or eating at night, adopting a sedentary lifestyle, and using cars for transportation most of the time

## Limitations

There is a limited number of studies exploring the relationship between mindful eating and diet quality as assessed by the KIDMED index among university students, which constitutes a limitation in terms of discussion and generalizability. In this study, data were collected using an electronic questionnaire. All measurements, including BMI, were based on self-reported data, and no retrospective dietary intake records were obtained. Future research could benefit from incorporating at least three-day food consumption records to more accurately assess dietary habits. Moreover, given that medical students receive coursework on nutrition and obesity, their responses may have been influenced by social desirability bias, potentially leading to overreporting of healthy behaviors. This factor should be considered when interpreting the findings. Additionally, the cross-sectional design of the study limits the ability to infer causality between mindful eating and dietary quality.

The KIDMED questionnaire, while practical for screening, is restricted to specific food categories and does not provide quantitative data. As such, it may yield incomplete results in evaluating overall adherence to the Mediterranean diet and alignment with national dietary recommendations. Future studies should incorporate comprehensive dietary assessment methods and further investigate the role of mindful eating in promoting diet quality.

Despite these limitations, the current findings offer valuable insights and may inform targeted interventions aimed at improving dietary behaviors and managing obesity risk among medical students.

## CONCLUSION

The association of mindful eating with low diet quality and obesity was not significant. First of all, healthy nutrition and interventions to reduce obesity should be implemented by considering risk groups. Intern physicians can be provided with access to healthier and free or easily affordable food on campus. Initiatives can be planned especially for male students to prepare cheap and easy meals.

## Supporting information

Title page

## Data Availability

All data produced in the present work are contained in the manuscript

file:///Users/ecem/Downloads/678939.pdf

