## Supplementary material for "Association between Mindful Eating and Obesity, Nutritional Status among 6th Grade Medical School Students": Title page

**ORCIDs:**

Penbe Ecem MISIRLIOĞLU: [**https://orcid.org/0000-0003-0944-4831**](https://orcid.org/0000-0003-0944-4831)

Hatice ŞİMŞEK: <https://orcid.org/0000-0001-7209-485X>

**Acknowledgements:** List all contributors who do not meet the criteria for authorship, such as technical assistants, writing assistants or head of department who provided only general support.

**Author(s) contribution(s):** The authors' contribution rate statement to this publication is as follows: Conception: P.E. M., H.Ş.; Design: P.E.M., H.Ş.; Supervision: H.Ş.; Data Collection and Processing: P.E.M., H.Ş.; Analysis-Interpretation: P.E.M., H.Ş.; Literature Review: P.E.M., Writing: P.E.M. Critical Review: P.E.M., H.Ş.

**Conflict of interest:** The authors declared no potential conflicts of interest with respect to the research, authorship, and/or publication of this article.

**Ethical approval**: All participants were informed about the aim and methods of the study, and signed an informed consent. The study was conducted in accordance with the Declaration of Helsinki, and the protocol was approved by the Ethics Committee of Non-Interventional Research Ethics Committee of Dokuz Eylul University (Date: 19/10/2020; Decision No:2020/26-35).

**Funding:** The authors received no financial support for the research, authorship, and/or publication of this article.
